# Refractive errors, ocular biometry and diabetic retinopathy: a systematic review

**DOI:** 10.1101/2020.02.17.20024000

**Authors:** Miao He, Haiying Chen, Wei Wang

## Abstract

Diabetic retinopathy (DR) is one of the major causes of visual impairment and blindness worldwide. The onset and progression of DR are influenced by systemic factors such as hyperglycemia and hypertension as well as ocular parameters. A better knowledge of the risk factors for DR is vital to improving the outcome of patients with DR and risk stratification. More recently, there has been increasing focus on the influence of myopia on DR development. Some observational studies have reported myopia being a protective factor for the development of DR, however the findings were inconsistent. In addition, it remains unclear whether it was myopia, axial length (AL), or other refractive factors that play the protective role. The protective mechanism against DR may be related to ocular elongation, posterior vitreous detachment, low perfusion in the retina and the abnormal cytokine profile. This systematic review will summarize the association of DR with refractory status as well as different refractive components including anterior chamber depth, refractory power of the lens, AL, and axial length-to-corneal radius ratio.

## 1. Introduction

While diabetic retinopathy (DR) is a common cause of blindness in middle-aged and elderly patients, its pathogensis remains unclear. Compared to the widely known systemic risk factors such as the duration of DR, glycemic control, blood pressure, research on ocular risk factors is limited.^1; 2; 3^ Myopia has emerged as a public challenge worldwide. In particular, high myopia increases the risk of blindness significantly due to its sight-threatening complications.^4^ It was estimated that half of the world’s population (5 billion) will develop myopia by 2050, and there will be one fifth of the population (1 billion) with high myopia.^5^ In some East Asian area, high myopia has become the leading cause of blindness.^6; 7^ The earliest study on the association between myopia and DR onset dated back to 1960.^8^ Previous studies have reported myopia as a protection factor for DR, however this was not widely accepted.^9; 10; 11^ Clinical data suggested that those with high myopia have a smaller risk for DR; in addition, it was less likely for diabetes patients with myopia to develop severe DR. Small hospital-based studies and epidemiology studies showed that myopia might be protective against DR, but the findings were inconsistent.^12; 13^ It remains unclear whether ocular structure, refractive parameters, or both contribute to this protective effect. Whether the protective effect of myopia against DR is related to the refractive status itself or ocular biometrics, including anterior chamber depth (ACD), corneal curvature, axial length (AL), axial length-to-corneal radius ratio (AL/CR ratio) is yet to be determined.

## 2. Refractive error and DR

Previous studies that investigated the relationship between refractive errors and DR were summerized in Table 1. In a clinical retrospective study of 19 participants with 38 eyes with asymmetric DR by Dogru and colleagues, there was zero case of proliferative diabetic retinopathy (PDR) in eyes with high myopia and complete posterior vitreous detachment (PVD).^14^ The group therefore proposed that high myopia and complete PVD may delay the progression of PDR. However, the study was based on a small sample size of 19 participants. Another study that evaluated 116 participants with anisometropia reported that after controlling for factors such as age, diabetes duration, ethnicity etc., high myopia could delay the onset of DR, and such protective effect was stronger with higher myopia.^15^ A number of population-based cross-sectional studies have investigated the relationship between refractive status and DR. A Singaporean study reported a reduced risk for DR, especially vision threatening DR (VTDR) in myopia.^12^ Another study in Korea, the Korea National Health and Nutrition Examination Survey (KNHANES), showed that with every 1 diopter increase in spherical equivalent (SE), there was a 30% higher incidence of DR.^16^ The Beijing Eye study also reported a 32% reduction in the risk for DR in myopia.^17^ In a population-based cohort study of 1210 young diabetic patients, myopia delayed the progression from DR to PDR after adjusting for other factors.^18^ On the other hand, the Singapore Indian Eye Study did not demonstrate a correlation between refractive error and DR, or between refractive error and VTDR.^19^ A more recent prospective cohort study also reached a similar conclusion that the SE and refractive error status were not associated with DR onset or progression.^20^ A common limitation of these studies is that there was no assessment of how myopia of different severity affects DR other than PDR.

**Table 1.**
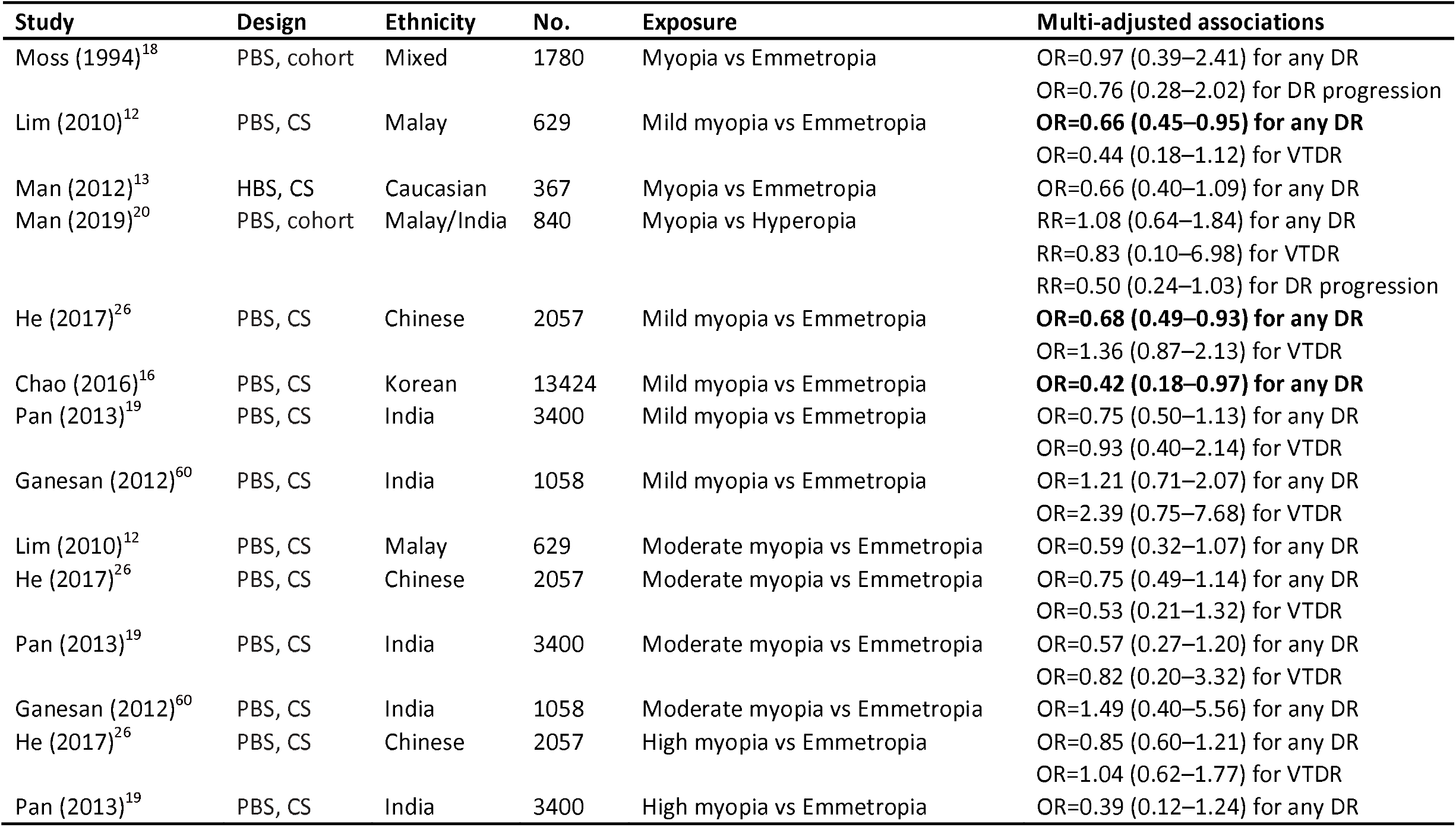
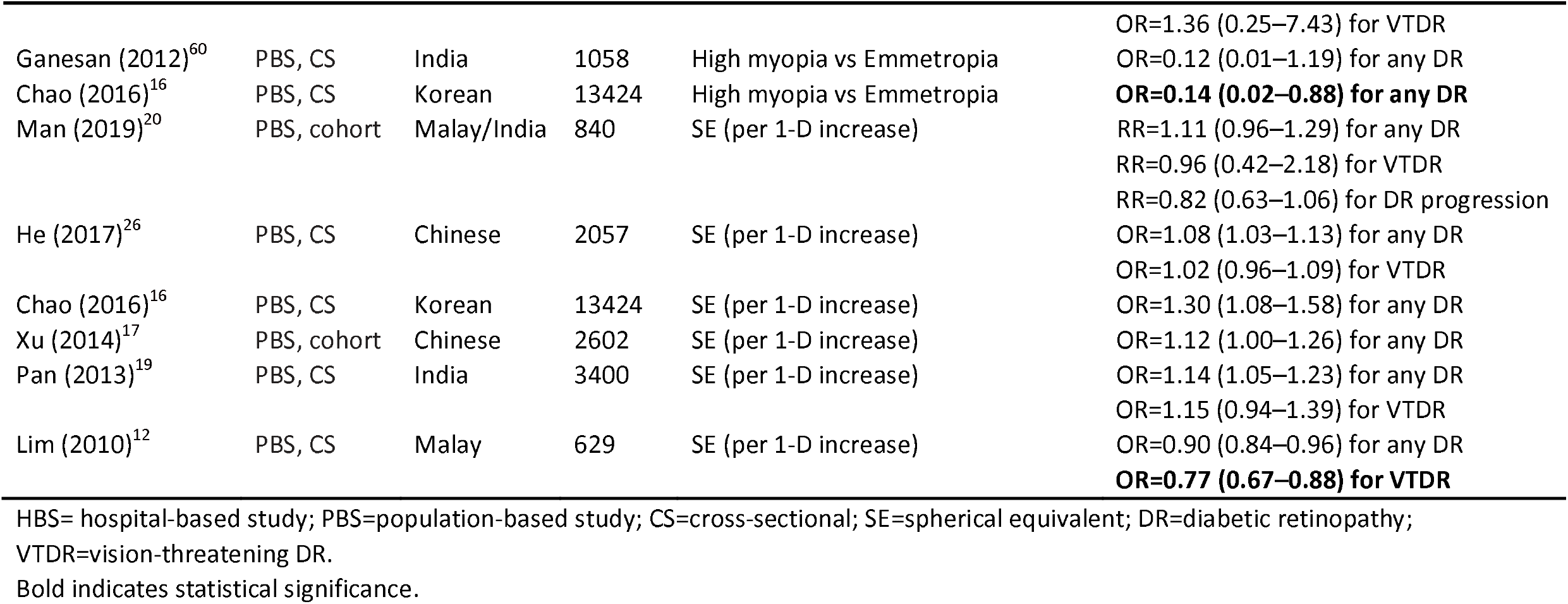
Previous studies investigating the influence of refractive errors on risk of diabetic retinopathy.

## 3. AL, axial length-to-corneal radius ratio and DR

Table 2 presents the relationship between AL, axial length-to-corneal radius (AL/CR) ratio and DR risk in different studies. A study that followed a group of Chinese patients for over 10 years reported that AL was negatively correlated with the development of PDR.^21^ In a study involving 630 eyes of 367 patients, Man et al.^13^ found that eyes with longer AL had lower risk for any DR and diabetic macular edema (DME), and that longer AL served as a protection factor for DR and DME. In a cohort with coexisting DME and cataract, the group with longer AL had better best corrected visual acuity (BCVA) than the control group at 12 months post vitrectomy.^22^ Another retrospective study on diabetes patients with anisometropia (interocular AL difference of > 1mm) showed that eyes with longer AL and thinner choroid had significantly lower incidence of PDR.^23^

**Table 2.** Summary of studies evaluating the associations of AL and AL/CR ratio with diabetic retinopathy.

| Study | Design | Ethnicity | No. | Exposure | Multi-adjusted associations |
| --- | --- | --- | --- | --- | --- |
| Jiang (2012)<sup>61</sup> | HBS, Case-control | Chinese | 168 | AL (per 1-mm increase) | OR=1.36 (0.98–1.88) for any DR |
| Yang (2012)<sup>21</sup> | HBS, Cross-sectional | Chinese | 166 | AL (per 1-mm increase) | OR=0.93 (0.71–1.21) for any DR<br>**OR=0.61 (0.39–0.94) for VTDR** |
| Man (2012)<sup>13</sup> | HBS, Cross-sectional | Caucasian | 367 | AL (per 1-mm increase) | **OR=0.76 (0.63–0.90) for any DR**<br>**OR=0.74 (0.62–0.89) for DME** |
| Xu (2014)<sup>17</sup> | PBS, Cohort, 10 yrs | Chinese | 2602 | AL (per 1-mm increase) | **OR=0.48 (0.33–0.71) for any DR** |
| Man (2019)<sup>20</sup> | PBS, Cohort, 6 yrs | Malay/India | 840 | AL (per 1-mm increase) | **RR=0.58 (0.38–0.88) for any DR**<br>RR=0.53 (0.11–2.70) for VTDR<br>RR=0.99 (0.55–1.80) for DR progression |
| He (2017)<sup>26</sup> | PBS, Cross-sectional | Chinese | 2057 | AL (per 1-mm increase)<br>AL/CR ratio | **OR=0.88 (0.81–0.95) for any DR**<br>OR=0.93 (0.83–1.05) for VTDR<br>**OR=0.43 (0.24–0.78) for any DR**<br>OR=0.72 (0.33–1.58) for VTDR |
| Pan (2013)<sup>19</sup> | PBS, Cross-sectional | India | 3400 | AL (per 1-mm increase) | **OR=0.73 (0.63–0.86) for any DR**<br>OR=0.73 (0.49–1.09) for VTDR |
| Wang (2019)<sup>24</sup> | PBS, Cross-sectional | Chinese | 1096 | AL (per 1-mm increase) | **OR=0.81 (0.70–0.95) for any DR** |
| Lim (2010)<sup>12</sup> | PBS, Cross-sectional | Malay | 629 | AL (per 1-mm increase) | **OR=0.86 (0.75–0.99) for any DR**<br>**OR=0.63 (0.40–0.99) for VTDR** |
| Bikbov (2019)<sup>27</sup> | PBS, Cross-sectional | Caucasian | 5899 | AL (per 1-mm increase) | P=0.26 for any DR |
| Ganesan (2012)<sup>60</sup> | PBS, Cross-sectional | India | 1058 | AL (Quartile 4 vs 1) | OR=0.90 (0.54–1.50) for any DR<br>OR=1.00 (0.31–3.22) for VTDR |

HBS= hospital-based study; PBS=population-based study; AL=axial length; DR=diabetic retinopathy; VTDR=vision-threatening DR; AL/CR= AL-to-corneal radius ratio; DME=diabetic macular edema. Bold indicates statistical significance.

Nevertheless, the relationship between AL and DR might vary amongst different ethnicities. In the Singapore Malay Eye Study (SiMES), every 1mm increase of AL was associated with a decreased risk for any DR, mild DR, and VTDR by 14%, 20% and 37%, respectively.^12^ Similarly, the recent Kailuan Eye Study in northern China reported that the odds of DR decreased by 19% for every 1mm increase in AL.^24^ The Beijing Eye Study also found that every 1mm increase in AL was associated with a reduced odds of DR by 38%.^25^ In the Singaporean Indian population, longer AL was negatively associated with any DR, but not with VTDR.^19^ A population study of 2057 type 2 diabetes mellitus (T2DM) adults in China reported that the refractive power of the lens, AL/CR ratio, ocular elongation may contribute to the protective role of AL against DR.^26^ Specifically, the group suggested that people with longer AL and bigger AL/CR ratio were less likely to develop DR. On the other hand, the Singapore Malay and Indian Eye Study which followed 840 diabetes patients for 6 years did not demonstrate a relationship between AL or other ocular biometric parameters and the risk of VTDR.^20^ The Ural Eye and Medical Study, a population-based study in Russia, also reported no correlation between AL and DR (P=0.16).^27^ Long AL is an indicator of axial myopia, and the rapidly expanding prevalence of myopia globally may have a potential impact on the incidence and prevalence of DR. More longitudinal studies involving various ethnicities are warranted.

## 4. ACD, lens parameters, corneal curvature and DR

There are fewer studies on the association between ACD, lens parameters and DR, and the results have been inconsistent (Table 3). Jiang and colleagues found that shallower ACD was associated with higher risk for DR onset and progression, as well as DME.^28^ In the Singapore Malay Eye Study (SiMES), every 1mm increase in ACD was associated with a reduced risk of middle DR and VTDR by 68% and 86%, respectively.^12^ On the other hand, the Beijing Eye Study and a hospital-based study in Australia found no association between ACD and DR.^13; 25^

**Table 3.** Relationship between the corneal curvature, anterior chamber depth, lens parameters, and diabetic retinopathy.

| Study | Design | Ethnicity | No. | Multi-adjusted associations |
| --- | --- | --- | --- | --- |
| **Corneal curvature** |  |  |  |  |
| Man (2019)<sup>20</sup> | PBS, cohort | Malay/India | 840 | RR=0.33 (0.11–1.01) for any DR<br>RR=0.12 (0.00–6.55) for VTDR<br>RR=0.65 (0.14–3.03) for DR progression |
| Man (2012)<sup>13</sup> | HBS, CS | Caucasian | 367 | OR=1.62 (0.76–3.48) for any DR |
| Jiang (2012)<sup>61</sup> | HBS, case-control | Chinese | 168 | OR=1.01 (0.98–1.04) for any DR |
| **Lens power** |  |  |  |  |
| He (2017)<sup>26</sup> | PBS, CS | Chinese | 2057 | OR=1.37 (0.99–1.89) for any DR<br>OR=1.01 (0.93–1.09) for VTDR |
| **Lens thickness** |  |  |  |  |
| Xu (2014)<sup>17</sup> | PBS, cohort | Chinese | 2602 | OR=1.23 (0.65–2.30) for any DR |
| Pierro (1996)<sup>29</sup> | HBS, CS | Caucasian | 57 | **P<0.05 for proliferative DR** |
| **Anterior chamber depth** |  |  |  |  |
| Man (2019) | PBS, cohort | Malay/India | 840 | RR=1.21 (0.63–2.34) for any DR<br>RR=0.31 (0.06–1.75) for VTDR<br>RR=1.85 (0.84–4.07) for DR progression |
| Xu (2014)<sup>17</sup> | PBS, cohort | Chinese | 2602 | OR=1.33 (0.94–1.88) for any DR |
| Jiang (2012)<sup>61</sup> | HBS, case-control | Chinese | 168 | OR=5.96 (1.78–3.81) for any DR |
| Lim (2010)<sup>12</sup> | HBS, CS | Malay | 629 | OR=0.73 (0.49–1.07) for any DR<br>**OR=0.14 (0.06–0.36) for VTDR** |

HBS= hospital-based study; PBS=population-based study; AL=axial length; DR=diabetic retinopathy; VTDR=vision-threatening DR.
Bold indicates statistical significance.

Studies on the relationship between lens parameters and DR have reached different conclusions. One study reported that the increase in lens thickness was associated with PDR in insulin-dependent diabetes patients.^29^ He and colleagues showed that higher refractive power of the lens was associated with a higher risk for DR, suggesting that the refractive power of the lens and other refractive factors may play a role in DR onset.^26^ However, Wiemer’s group did not demonstrate any association between lens parameters and DR after adjusting for confounding factors.^30^ The inconsistent findings might be attributable to the differences in research methodology. As such, the influence of ACD and lens parameters on DR warrants further population-based studies of a large sample size.

## 5. Fundus tessellation, peripapillary atrophy and DR

Fundus tessellation is darkly pigmented choroid in the posterior retina, which is commonly seen in eyes with myopia or age-related degeneration, and is an important consideration in the diagnosis of retinal-choroidal degeneration. Peripapillary atrophy (PPA), also known as disc crescent, is closely associated with myopia.^31; 32; 33^ Fundus tessellation and PPA, the two common early signs of axial high myopia, are present in 81%-98% and 90%-93% of highly myopic eyes, respectively.^34; 35^ The prevalence of PPA is 15%-20% in the general population, and can be as high as 67% in some reports.^36; 37^ Only one study so far has investigated the correlation between DR and these two signs. A cross-sectional analysis of 592 diabetes patients in the Singapore Chinese Eye Study showed that the presence of PPA and fundus tessellation in diabetic patients was correlated with a lower risk for DR, independent of other diabetes risk factors, AL, or refractive error.^38^

## 6. Possible mechanisms of the protective effect

Based on previous studies discussed in the earlier part of this report, myopic refractive error and long AL may be protective factors against DR. Possible mechanisms might be attributable to the regulating effect of the increase in AL, posterior vitreous detachment, low blood flow perfusion in the retina, choroidal thinning, and the alteration of cytokine profile.^39; 40; 41; 42; 43; 44^ While the exact mechanisms of how myopia is protective against DR remain elusive, most of the attention is placed on the pathological changes of the eyeball incurred by the increasing AL during the progression of myopia. ^39; 40; 41^

### 6.1 Decrease in blood flow volume

One theory proposes that as myopia progresses, the scleral wall is stretched, resulting in distortion of the posterior retina and a reduction in blood flow in the retina.^45^ The reduction in blood flow volume would decrease the pressure to vessel wall, which may reduce the risk for DR onset. This is consistent with the theory that the increased retinal blood flow volume is a significant risk factor for non-PDR developing into PDR, as the retinal blood flow volume is greater in the diabetes group compared to the non-diabetes group.^45^ Nevertheless, one study that used Heidelberg Retinal Flowmeter to measure the blood flow in the capillaries did not demonstrate the reduction of retinal capillary blood flow as a main protective factor against DR.^40^

### 6.2 Change of blood flow dynamics

In high myopia, there are varying degrees of atrophy and degeneration in the retina and choroid. As AL increases, retinal and choroidal tissues become thinner and vascular diameter reduces.^49; 50^ This results in an increase in the trans-vessel pressure and a decrease in blood flow in the retina. The reduction in retinal blood flow perfusion not only reduces the physical stress to the retinal vascular wall, but also dampens the biochemical damage of hyperglycemia in diabetes. Patients with high myopia are more likely to develop aged-related degeneration in the retinal microvascular beds before 40 years of age, whereas retinal vascularture degeneration usually occurs in T2DM patients after 40. It is possible that retinal and choroidal degeneration occurs earlier on in patients with T2DM and high myopia, leading to lower retinal blood flow perfusion.^3; 38^ This may partially offset the damage from high blood flow perfusion that tends to occur in DR. In addition, intraocular pressure (IOP) of myopic patients increases as myopia progresses, and IOP of patients with moderate myopia is higher compared to IOP of people without myopia.^51^ Therefore, with the same systolic blood pressure, perfusion pressure of the eye decreases as IOP increases, which would to some extent influence the hemodynamics in the retina, hence the onset and progression of DR in highly myopic patients.

### 6.3 Decrease in oxygen demand

In myopia, the demand of oxygen decreases as the posterior scleral extends and the retina thins, which may to some extent alleviate the effect of hypoxia on the retina due to diabetes. Jonas et al observed a reduction in vascular endothelial growth factor (VEGF) concentration in the vitreous humor in eyes with long AL, an indication of relatively lighter hypoxic effect on the retina.^52^ Man and collegues analyzed the multifocal macular electroretinogram (fm-ERG) and oxygen consumption in retinal veins and arteries in a cohort of 50 healthy participants, and found that artery-vein oxygen consumption and retinal function decreased as AL increased.^39^ Hence, the reduction in retinal oxygen consumption due to long AL may reduce the hypoxic stress and the risk of DR in diabetes patients. In addition, there is reduced oxygen demand when the function of the outer retina deteriorates in high myopia, and the hypoxic stress on the retina is relieved as more oxygen is obtained from the thinner choroid, which together may delay the onset or progression of DR. Therefore, due to the reduced metabolism in the atrophied choroid and outer retina as well as by enabling easier oxygen diffusion through the thinner retinal and choroidal tissues, high myopia could serve as a protective factor against DR.

### 6.4 Posterior vitreous detachment

Posterior vitreous detachment (PVD) occurs when AL increases but the vitreous body fails to extend with the globe in highly myopic eyes. PVD and liquefaction of the vitreous humor are common in highly myopic eyes, which disables the supporting structure for neovascularization and allows for more oxygen absorption through the liquefied vitreous humor and aqueous fluid.^53^ As a result, hypoxia in the retina is partially relieved and the micro-environment for neovascularization is altered. Other studies also reported that the fiber scaffolds for neovascular proliferation in the vitreous body disappeared and oxygen diffused more easily in the liquefied vitreous humor in complete PVD, hence delaying the progression of NPDR to neovascularization and PDR. Evidence also suggested that DR patients with complete PVD had a significantly lower risk for neovascularization in the retina and the optic disc. ^54; 55^

### 6.5 Change in cytokine profile

Many cytokines are implicated in the pathogenesis of DR such as endothelial injury, retinal proliferation and neovascularization. Dysregulation of VEGF, pigment epithelium derived factor (PEDF), tumor necrosis factor (TNF), erythropoietin (EPO), transforming growth factor beta (TFG-β), endothelin (ET-1) have been implicated in both DR and high myopia.^44; 52; 56; 57; 58^ The altered expression of these cytokines in myopia may have an effect on DR progression. A few studies recorded lower concentrations of VEGF and EPO in the vitreous in highly myopic eyes, and proposed that the reduced expression of VEGF and EPO might help to retard the DR progression in highly myopic eyes.^42^ Another study found that the PEDF concentration was lower in patients with high myopia than in control subjects, which might be due to the degeneration of retinal ganglion cells and retinal pigment epithelium that produce the PEDF.^59^ In patients with diabetes, high myopia may have an accommodative effect by altering the expression profile of certain cytokines which are implicated in the onset and progression of DR.

## 7. Conclusion

There is increasing evidence suggesting that high myopia may be a protective factor against the onset and progression of DR. However, due to the differences in study methods as well as quality and statistical analyses, results from previous studies have been inconsistent. To date, no consensus has been reached regarding the association between myopia and DR. Robust evidence is lacking to elucidate the association between AL, ACD, lens parameters, and the risk for DR. Many studies did not account for confounding factors such as age, gender and glycemic control. Whether or not retinal changes due to high myopia predate DR remains unclear. We also suggest future research look into the underlying choroidal and retinal changes in high myopia, and their implications on DR, as opposed to merely the association between ocular biometrics in myopia and DR. DR is a complex pathological process where multiple factors contribute collectively. Current studies suggest that the protecting effect of myopia against DR could be related to the prolongation of AL, posterior vitreous detachment, low retinal blood flow perfusion, choroidal thinning and the altered expression of cytokines. The exact mechanisms of how myopia, AL and ocular biometrics influence the onset and progression of DR are yet to be determined. Further investigation into the protective and risk factors of DR will provide valuable basis for potential new intervention and treatment options for DR.

## 8. Methods of Literature Search

Systematic search was conducted in electronic databases including Pubmed (http://www.ncbi.nlm.nih.gov/pubmed) and Embase.com (http://www.embase.com). The ARVO Meeting Abstracts (http://www.iovs.org/search.dtl?arvomtgsearch=true) were retrieved for additional studies. The following key words were used in the search: myopia, refractive error, short sight, diabetic retinopathy, diabetic eye disease, axial length, anterior chamber depth, corneal curvature, corneal radius, lens thickness, lens power, refractive power of lens. All abstracts were reviewed and appropriate studies were obtained in full for complete review. The references of included studies and related reviews were screened for additional articles. Articles wrriten in English were included, and those written in languages other than English were considered when English abstracts were available.

## Data Availability

All data were included in the manuscript.

## Acknowledgments

The authors declare no conflict of interest with respect to the topic and the content of this manuscript. This study was supported by the National Natural Science Foundation of China (81900866).

## References

1. Leasher JL, Bourne RR, Flaxman SR, et al.: Global Estimates on the Number of People Blind or Visually Impaired by Diabetic Retinopathy: A Meta-analysis From 1990 to 2010. Diabetes Care 39:1643–1649, 2016

2. Yau JW, Rogers SL, Kawasaki R, et al.: Global prevalence and major risk factors of diabetic retinopathy. Diabetes Care 35:556–564, 2012

3. Stitt AW, Curtis TM, Chen M, et al.: The progress in understanding and treatment of diabetic retinopathy. Prog Retin Eye Res 51:156–186, 2016

4. Morgan IG, Ohno-Matsui K, Saw SM: Myopia. Lancet 379:1739–1748, 2012

5. Holden BA, Fricke TR, Wilson DA, et al.: Global Prevalence of Myopia and High Myopia and Temporal Trends from 2000 through 2050. Ophthalmology 123:1036–1042, 2016

6. Liu HH, Xu L, Wang YX, et al.: Prevalence and progression of myopic retinopathy in Chinese adults: the Beijing Eye Study. Ophthalmology 117:1763–1768, 2010

7. Wong YL, Sabanayagam C, Ding Y, et al.: Prevalence, Risk Factors, and Impact of Myopic Macular Degeneration on Visual Impairment and Functioning Among Adults in Singapore. Invest Ophthalmol Vis Sci 59:4603–4613, 2018

8. Jain IS, Luthra CL, Das T: Diabetic retinopathy and its relation to errors of refraction. Arch Ophthalmol 77:59–60, 1967

9. Wang X, Tang L, Gao L, et al.: Myopia and diabetic retinopathy: A systematic review and meta-analysis. Diabetes Res Clin Pract 111:1–9, 2016

10. Fu Y, Geng D, Liu H, Che H: Myopia and/or longer axial length are protective against diabetic retinopathy: a meta-analysis. Acta Ophthalmol 94:346–352, 2016

11. Man RE, Sasongko MB, Wang JJ, Lamoureux EL: Association between myopia and diabetic retinopathy: a review of observational findings and potential mechanisms. Clin Exp Ophthalmol 41:293–301, 2013

12. Lim LS, Lamoureux E, Saw SM, et al.: Are myopic eyes less likely to have diabetic retinopathy? Ophthalmology 117:524–530, 2010

13. Man RE, Sasongko MB, Sanmugasundram S, et al.: Longer axial length is protective of diabetic retinopathy and macular edema. Ophthalmology 119:1754–1759, 2012

14. Dogru M, Inoue M, Nakamura M, Yamamoto M: Modifying factors related to asymmetric diabetic retinopathy. Eye (Lond) 12 (Pt 6):929–933, 1998

15. Bazzazi N, Akbarzadeh S, Yavarikia M, et al.: HIGH MYOPIA AND DIABETIC RETINOPATHY: A Contralateral Eye Study in Diabetic Patients With High Myopic Anisometropia. Retina 37:1270–1276, 2017

16. Chao DL, Lin SC, Chen R, Lin SC: Myopia is Inversely Associated With the Prevalence of Diabetic Retinopathy in the South Korean Population. Am J Ophthalmol 172:39–44, 2016

17. Xu J, Xu L, Wang YX, et al.: Ten-year cumulative incidence of diabetic retinopathy. The Beijing Eye Study 2001/2011. PLoS One 9:e111320, 2014

18. Moss SE, Klein R, Klein BE: Ocular factors in the incidence and progression of diabetic retinopathy. Ophthalmology 101:77–83, 1994

19. Pan CW, Cheung CY, Aung T, et al.: Differential associations of myopia with major age-related eye diseases: the Singapore Indian Eye Study. Ophthalmology 120:284–291, 2013

20. Man R, Gan A, Gupta P, et al.: Is Myopia Associated with the Incidence and Progression of Diabetic Retinopathy? Am J Ophthalmol 208:226–233, 2019

21. Yang KJ, Sun CC, Ku WC, et al.: Axial length and proliferative diabetic retinopathy. Optom Vis Sci 89:465–470, 2012

22. Wakabayashi Y, Kimura K, Muramatsu D, et al.: Axial length as a factor associated with visual outcome after vitrectomy for diabetic macular edema. Invest Ophthalmol Vis Sci 54:6834–6840, 2013

23. Kim DY, Song JH, Kim YJ, et al.: Asymmetric diabetic retinopathy progression in patients with axial anisometropia. Retina 38:1809–1815, 2018

24. Wang Q, Wang YX, Wu SL, et al.: Ocular Axial Length and Diabetic Retinopathy: The Kailuan Eye Study. Invest Ophthalmol Vis Sci 60:3689–3695, 2019

25. Xu L, Cao WF, Wang YX, et al.: Anterior chamber depth and chamber angle and their associations with ocular and general parameters: the Beijing Eye Study. Am J Ophthalmol 145:929–936, 2008

26. He J, Xu X, Zhu J, et al.: Lens Power, Axial Length-to-Corneal Radius Ratio, and Association with Diabetic Retinopathy in the Adult Population with Type 2 Diabetes. Ophthalmology 124:326–335, 2017

27. Bikbov MM, Kazakbaeva GM, Gilmanshin TR, et al.: Axial length and its associations in a Russian population: The Ural Eye and Medical Study. PLoS One 14:e211186, 2019

28. Jiang JJ, Li XX, Yuan L, et al.: Ocular biological structures and relevant risk factors in the occurrence of diabetic retinopathy in diabetes mellitus patients. Zhonghua Yan Ke Za Zhi 48:898–902, 2012

29. Pierro L, Brancato R, Zaganelli E, et al.: Correlation of lens thickness with blood glucose control in diabetes mellitus. Acta Ophthalmol Scand 74:539–541, 1996

30. Wiemer NG, Dubbelman M, Kostense PJ, et al.: The influence of diabetes mellitus type 1 and 2 on the thickness, shape, and equivalent refractive index of the human crystalline lens. Ophthalmology 115:1679–1686, 2008

31. Ruiz-Medrano J, Montero JA, Flores-Moreno I, et al.: Myopic maculopathy: Current status and proposal for a new classification and grading system (ATN). Prog Retin Eye Res 69:80–115, 2019

32. Yokoi T, Jonas JB, Shimada N, et al.: Peripapillary Diffuse Chorioretinal Atrophy in Children as a Sign of Eventual Pathologic Myopia in Adults. Ophthalmology 123:1783–1787, 2016

33. Ohno-Matsui K, Kawasaki R, Jonas JB, et al.: International photographic classification and grading system for myopic maculopathy. Am J Ophthalmol 159:877–883, 2015

34. Yan YN, Wang YX, Xu L, et al.: Fundus Tessellation: Prevalence and Associated Factors: The Beijing Eye Study 2011. Ophthalmology 122:1873–1880, 2015

35. Guo Y, Liu L, Zheng D, et al.: Prevalence and Associations of Fundus Tessellation Among Junior Students From Greater Beijing. Invest Ophthalmol Vis Sci 60:4033–4040, 2019

36. Zhang Q, Wang YX, Wei WB, et al.: Parapapillary Beta Zone and Gamma Zone in a Healthy Population: The Beijing Eye Study 2011. Invest Ophthalmol Vis Sci 59:3320–3329, 2018

37. Skaat A, De Moraes CG, Bowd C, et al.: African Descent and Glaucoma Evaluation Study (ADAGES): Racial Differences in Optic Disc Hemorrhage and Beta-Zone Parapapillary Atrophy. Ophthalmology 123:1476–1483, 2016

38. Tan N, Tham YC, Ding Y, et al.: Associations of Peripapillary Atrophy and Fundus Tessellation with Diabetic Retinopathy. Ophthalmol Retina 2:574–581, 2018

39. Man RE, Lamoureux EL, Taouk Y, et al.: Axial length, retinal function, and oxygen consumption: a potential mechanism for a lower risk of diabetic retinopathy in longer eyes. Invest Ophthalmol Vis Sci 54:7691–7698, 2013

40. Man RE, Sasongko MB, Xie J, et al.: Decreased retinal capillary flow is not a mediator of the protective myopia-diabetic retinopathy relationship. Invest Ophthalmol Vis Sci 55:6901–6907, 2014

41. Bartol-Puyal FA, Isanta C, Ruiz-Moreno O, et al.: Distribution of Choroidal Thinning in High Myopia, Diabetes Mellitus, and Aging: A Swept-Source OCT Study. J Ophthalmol 2019:3567813, 2019

42. Kwon SH, Shin JP, Kim IT, Park DH: Aqueous Levels of Angiopoietin-like 4 and Semaphorin 3E Correlate with Nonperfusion Area and Macular Volume in Diabetic Retinopathy. Ophthalmology 122:968–975, 2015

43. Finzi A, Cellini M, Strobbe E, Campos EC: ET-1 plasma levels, choroidal thickness and multifocal electroretinogram in retinitis pigmentosa. Life Sci 118:386–390, 2014

44. .Jain A, Saxena S, Khanna VK, et al.: Status of serum VEGF and ICAM-1 and its association with external limiting membrane and inner segment-outer segment junction disruption in type 2 diabetes mellitus. Mol Vis 19:1760–1768, 2013

45. Ohno-Matsui K, Jonas JB: Posterior staphyloma in pathologic myopia. Prog Retin Eye Res 70:99–109, 2019

46. Sun Z, Tang F, Wong R, et al.: OCT Angiography Metrics Predict Progression of Diabetic Retinopathy and Development of Diabetic Macular Edema: A Prospective Study. Ophthalmology 126:1675–1684, 2019

47. Hwang TS, Gao SS, Liu L, et al.: Automated Quantification of Capillary Nonperfusion Using Optical Coherence Tomography Angiography in Diabetic Retinopathy. JAMA Ophthalmol 134:367–373, 2016

48. Samara WA, Shahlaee A, Adam MK, et al.: Quantification of Diabetic Macular Ischemia Using Optical Coherence Tomography Angiography and Its Relationship with Visual Acuity. Ophthalmology 124:235–244, 2017

49. Li M, Yang Y, Jiang H, et al.: Retinal Microvascular Network and Microcirculation Assessments in High Myopia. Am J Ophthalmol 174:56–67, 2017

50. Yang Y, Wang J, Jiang H, et al.: Retinal Microvasculature Alteration in High Myopia. Invest Ophthalmol Vis Sci 57:6020–6030, 2016

51. Jonas JB, Nagaoka N, Fang YX, et al.: Intraocular Pressure and Glaucomatous Optic Neuropathy in High Myopia. Invest Ophthalmol Vis Sci 58:5897–5906, 2017

52. Jonas JB, Tao Y, Neumaier M, Findeisen P: VEGF and refractive error. Ophthalmology 117:2231–2234, 2010

53. Akiba J: Prevalence of posterior vitreous detachment in high myopia. Ophthalmology 100:1384–1388, 1993

54. Anderson W, Piggott K, Bao YK, et al.: Complete Posterior Vitreous Detachment Reduces the Need for Treatment of Diabetic Macular Edema. Ophthalmic Surg Lasers Imaging Retina 50:e266–e273, 2019

55. Gella L, Raman R, Kulothungan V, Sharma T: Prevalence of posterior vitreous detachment in the population with type II diabetes mellitus and its effect on diabetic retinopathy: Sankara Nethralaya Diabetic Retinopathy Epidemiology and Molecular Genetic Study SN-DREAMS report no. 23. Jpn J Ophthalmol 56:262–267, 2012

56. Jonas JB, Jonas RA, Neumaier M, Findeisen P: Cytokine concentration in aqueous humor of eyes with diabetic macular edema. Retina 32:2150–2157, 2012

57. Zhu D, Yang DY, Guo YY, et al.: Intracameral interleukin 1beta, 6, 8, 10, 12p, tumor necrosis factor alpha and vascular endothelial growth factor and axial length in patients with cataract. PLoS One 10:e117777, 2015

58. Chen W, Guan Y, He G, et al.: Aqueous Levels of Pigment Epithelium-Derived Factor and Macular Choroidal Thickness in High Myopia. J Ophthalmol 2015:731461, 2015

59. Ogata N, Imaizumi M, Miyashiro M, et al.: Low levels of pigment epithelium-derived factor in highly myopic eyes with chorioretinal atrophy. Am J Ophthalmol 140:937–939, 2005

60. Ganesan S, Raman R, Reddy S, et al.: Prevalence of myopia and its association with diabetic retinopathy in subjects with type II diabetes mellitus: A population-based study. Oman J Ophthalmol 5:91–96, 2012

61. Jiang J, Li X, Yuan L, et al.: Ocular biological structures and relevant risk factors in the occurrence of diabetic retinopathy in diabetes mellitus patients. Chinese Journal of Ophthalmology 48:898–902, 2012

